# Peripheral signature of altered synaptic integrity in young onset Cannabis Use Disorder: A proteomic study of circulating extracellular vesicles

**DOI:** 10.1101/2022.06.17.22276563

**Authors:** Suhas Ganesh, TuKiet T. Lam, Rolando Garcia-Milian, Deepak D’Souza, Angus C. Nairn, Katya Elgert, Erez Eitan, Mohini Ranganathan

## Abstract

**Background:** The prevalence of cannabis use and Cannabis Use Disorder (CUD) are highest amongst adolescents and young adults. A lack of brain tissues from patients with CUD limits the ability to examine the molecular basis of cannabis related neuropathology. Proteomic studies of neuron-derived extracellular vesicles (NDEs) isolated from the biofluids may reveal markers of neuropathology in CUD.

**Methods:** NDEs were extracted using ExoSORT, an immunoaffinity method, from plasma samples of 10 patients with young onset CUD and 10 matched controls. Differential proteomic profiles of NDEs between groups was explored with Label Free Quantification (LFQ) mass spectrometry. Selected differentially abundant proteins were validated using orthogonal methods.

**Results:** A total of 231 (+/- 10) unique proteins were identified in NDE preparations of which 28 were differentially abundant between groups. The difference in abundance properdin, encoded by the *CFP* gene surpassed the significance threshold after false discovery rate correction.

Notably, SHANK1 (SH3 and multiple ankyrin repeat domains protein 1), an adapter protein at the post-synaptic density, was found to be depleted in the CUD compared to control NDE preparations.

**Discussion:** The study shows that LFQ mass spectrometry proteomic analysis of NDEs derived from plasma may yield important insights into the synaptic pathology associated with CUD. Optimization of this approach may lead to a novel assay to study altered proteomic signalling in the brain using liquid biopsy in diverse neuropsychiatric syndromes.

## Introduction

Cannabis is one of the most commonly used drugs in the United Sates. The prevalence of cannabis use (∼33%) and Cannabis Use Disorder (CUD) (∼6%) are highest amongst adolescents and young adults ^1, 2^. CUD is associated with several persistent neuropsychiatric and cognitive consequences, particularly with earlier onset and heavier use ^3, 4^. Preclinical data demonstrate neuronal/synaptic pathology ^5-12^ and immunomodulatory changes (microglial and astrocytic activation) with repeated cannabinoid exposure ^13^. In humans, challenges in obtaining brain tissues, especially amongst young adults early in the course of CUD, limits the ability to examine the molecular basis of cannabis related neuropathology.

Extracellular Vesicles (EVs), secreted from various tissues can be assayed from biological fluids ^14^. In the central nervous system (CNS), EVs are secreted by both neuronal and glial cells and may be involved in processes such as intercellular communication, immune responses, and synaptic plasticity. EVs cross the blood brain barrier carrying CNS lipids, protein and RNA, making them attractive as reservoirs for biomarker discovery^15^. Relevant to the current investigation, EV membranes are enriched for endocannabinoids and activate cannabinoid receptors (CB1R). Thus specific EVs may play a role in endocannabinoid signalling, and may thus be sensitive to chronic cannabis exposure ^16^.

Advances in mass spectrometry (MS) approaches allow profiling proteomes in body-fluids (e.g., plasma) or the sub-proteomes within organelles of interest (e.g., EVs) ^17^. Label Free Quantitative (LFQ) proteomics approach permits detection and relative quantification of a large number of proteins, and thus can help in identifying pathway level perturbations in protein abundance. Recent studies have used MS approaches to evaluate EV proteomes relevant to disease states ^17-20^.

Few studies to date have used MS-based proteomics to evaluate the proteome of circulating EVs of neuronal origin - referred henceforth as Neuron Derived Extracellular Vesicles (NDE). There is currently little known about optimal methods of sample processing, enrichment and proteomic analysis approaches for plasma NDEs. In this study, we first aimed to develop protocols to optimize the methods for MS proteomic analysis of plasma-derived NDE. Next, we aimed to examine the effects of recurrent cannabis exposure during adolescence and young adulthood, on the proteomic signatures in circulating NDEs. Finally, we examined the profile of the EV protein cargo and its relevance to the neuropathology of CUD, with a focus on examining the differential abundance of synaptic proteins identified with an optimized LFQ proteomics approach.

## Methods

### Assay optimization

Pooled plasma samples, n = 4 of 0.5 ml each were obtained from a commercially available pooled plasma source (BioIVT, HMN699478 and HMN699479) to optimize methods of NDE extraction for proteomic profiling.

EVs were extracted from 0.5 ml plasma at NeuroDex (Natick, MA) using a standard EV isolation kit (4478360, Invitrogen; Thermo Fisher Scientific, Waltham, MA) followed by 0.2-micron filtration for total EV and ExoSORTTM (NDX00121, NeuroDex, Natick MA). Support for this new kit can be found in Supplemental Figure 1 and patent application (PCT - WO 2022/058881 A1). As residual contamination with free plasma proteins is a concern during label-free MS proteomics, we tested 4 different approaches of plasma NDE preparation to determine an optimal method - 1. M1 - Standard Exosort™ NDE extraction; 2. M2 - Standard Exosort™ NDE extraction + plasma protein depletion (High Select™ HSA/Immunoglobulin Depletion Mini Spin Columns, A36365, Thermofisher Scientific, Waltham, MA); 3. M3 - Erythrocyte derived EVs (CD235A, glycophorin A) depletion followed by Exosort™ NDE extraction; 4. M4 - Erythrocyte derived EVs depletion (CD235A, glycophorin A) followed by Exosort™ NDE extraction + plasma protein depletion (High Select™ HSA/Immunoglobulin Depletion Mini Spin Columns, A36365, Thermofisher Scientific, Waltham, MA). For NDE preparations in CUD and matched healthy controls, the best performing protocol from the above four methods was chosen for MS proteomics.

### Electron microscopy

Following ExoSORT, NDEs were resuspended in 3% PFA and visualized by negative staining (Uranyl Acetate) using a Morgagni transmission electron microscope (FEI, Hillsboro, OR), operating at 80 kV and equipped with a Nanosprint5 CMOS camera (AMT, Woburn, MA). Experiments were preformed with the kind help of Dr. Berith Isaaks (Brandeis University Cell Imaging facility) (Supplemental Figure 1).

### Nanoparticle tracking analysis

Following ExoSORT, NDEs were diluted with pre-filtered PBS (20 mm filters), and NTA analysis was performed using Nanosight500 (Malvern Panalytical) as described previously^21^ (Supplemental Figure 1).

### Label Free Quantification

#### Sample Preparation

NDEs in their respective extraction buffer were vortexed then centrifuged at 14,600 RPM at 4°C for 10 minutes. An aliquot of 175 µL of the supernatant was taken and proteins were precipitated utilizing a methanol/chloroform/water precipitation procedure ^22^. The resulting pellet was dissolved in 20 µL of 8M urea/0.4M ammonium bicarbonate, reduced with 2 µL of 45 mM dithiothreitol (DTT) at 37°C for 30 minutes, and subsequently alkylated with 2 µL of 100 mM iodoacetamide at room temperature for 30 minutes in the dark. The solution was diluted with 51 µL water and digested with 5 µL of 0.1 µg/µL LysC at 37°C overnight followed by 1µL of 0.5 µg/µL trypsin at 37°C for 7 hours. The digestion was quenched by acidifying with 4 µL of 20% trifluoro acetic acid. The peptide solution was then desalted using a mini RP C18 desalting columns (The Nest Group, Ipwich, MA). Eluted peptides were dried in a speedvac, and stored at -80C until data collection.

#### Data Collection

The proteomic profiling of the NDEs was performed at the Discovery Core of the Yale/NIDA Neuroproteomics Center, Yale University School of Medicine, New Haven, United States. Due to the paucity of published studies that enumerate the proteomic profile of circulating NDEs, specifically in the context of diseases involving brain pathology such as addiction, in this study, we employed a discovery proteomics approach. LFQ data-dependent acquisition (DDA) was performed on a Thermo Scientific Q-Exactive HFX mass spectrometer connected to a Waters M-Class ACQUITY UPLC system equipped with a Waters Symmetry® C18 180 μm × 20 mm trap column and a 1.7-μm, 75 μm × 250 mm nanoACQUITY UPLC column (35°C). 5 µl of each digest were reconstituted in Buffer A (0.1% FA in water) to a 0.05 µg/µl concentration and injected in block randomized order. UPLC peptide trapping was carried out for 3 min at 5 µl/min in 99% Buffer A and 1% Buffer B [(0.075% FA in acetonitrile (ACN)] prior to eluting with linear gradients that reached 6% B at 2 min, 25% B at 200 min, and 85% B at 205 min. Two blanks (1st 100% ACN, 2nd Buffer A) followed each injection to ensure against sample carry over. Settings for the Q-Exactive HFX mass spectrometer include: 45,000 MS scan resolution with AGC target of 3e6 (Max IT of 100ms) and scan range of 200-2000 m/z in profile mode; 15,000 MS^2^ scan resolution with AGC target of 1e5 (Max IT of 50ms) and scan range of 200-2000; and Top20 peptide HCD fragmentation consist of isolation window of 1.6 m/z, normalized collision energy of 28, preference for 2+ charge state, and dynamic exclusion of 20.0 seconds. All MS and MS/MS peaks were detected in the Orbitrap.

The initial protocol optimization phase for LC-MS/MS data was processed with Proteome Discoverer Software (v2.2, ThermoFisher Scientific, Waltham, MA) with protein identification carried out using an in-house Mascot search engine (v2.7, Matrix Science, Boston, MA) and the Swiss Protein database () with taxonomy restricted to H. sapiens. Carbamidomethyl (Cys) was a fixed modification and oxidation of Methionine (Met) was a variable modification. Two missed tryptic cleavages were allowed, precursor mass tolerance was set to 10 ppm, and fragment mass tolerance was set to 0.02 Da. The significance threshold was set based on a False Discovery Rate (FDR) of 5%, and MASCOT peptide score of >95% confidence. Subsequent LFQ data were processed with Progenesis QI software (v4.2, Waters Inc., Milford, MA) with similar Mascot search parameters as in the development part. Similar Progenesis QI and

Mascot search parameters are found in Franzen et al (2020) ^23^, and with protein requirements of at least 1 unique peptide with peptide score >30 (e.g. 95% confidence).

### Case-control analysis

For the analysis on the impact of chronic cannabis exposure on the proteomic profile of NDE, plasma samples were obtained from individuals with young onset CUD and age and gender matched healthy controls. Young onset was defined as initiation of regular cannabis use before the age of 18 years.

### Validation of differentially abundant proteins

Three of the most differentially abundant proteins in NDE from CUD and matched controls were validated with Enzyme Linked Immunosorbent Assay (ELISA) using commercially available antibodies (Thermo Scientific, Cat. EH383RB, Sigma-Aldrich, Cat. HCMP2MAG-19K-07, MyBioSource Cat. MBS9339587). All assays were run in accordance with the manufacturer instructions. ELISA plates were read using CLARIOstar plate reader and Luminex assays with Luminex 200 instrument.

### Analysis approach

Enrichment of the list of proteins identified by LFQ analysis was examined using the PANTHER (Protein ANalysis THrough Evolutionary Relationships) Classification System ^24^ and a Fisher’s exact test with false discovery rate (FDR) correction of p < 0.05. The sensitivity of the purification approaches to identify proteins predominantly expressed in brain compared to other tissues, was examined with enrichment for ‘brain-enriched’ and ‘brain-elevated’ proteins as defined in the Human Brain Proteome database, using a Fisher’s exact test. The protein probabilities, spectral counts, and unique peptide counts for ‘brain-enriched’ and ‘brain-elevated’ proteins in LFQ analysis were compared using violin plots. Among the four purification approaches, the one resulting in the best yield of proteins across these parameters was selected for NDE preparation to compare the differential protein abundance in CUD and controls.

Differential protein abundances between CUD and controls were measured as fold changes in mean relative protein abundance, with an FDR cut off <0.1. Proteins that have been previously identified in the processes of synaptic physiology, neuroinflammation and astrocytic reaction were specifically investigated in the present study. In R version 3.6.1 ^25^, tidyverse ^26^ and ggpubr ^27^ packages were employed for data analysis and plotting. Pathway analysis was performed using Ingenuity Pathway Analysis (Qiagen, Redwood City, CA). Differentially abundant proteins were mapped to corresponding IPA identifier and used in the overrepresentation analysis.

Significant pathways are those with FDR p <0.05 of the Fisher’s exact test. Blue bars correspond to downregulated pathways (z-score < 0).

## Results

### A. Protocol optimization

A total of 465 unique proteins were identified by the Mascot search algorithm across the four different NDE preparations from pooled plasma replicates. The top 20 enriched GO ‘cellular component’ terms derived from an overrepresentation analysis against a background referece list of human protein coding genes (n =20595, PANTHER database) are presented in Table 1. This analysis confirmed a broad enrichment profile of NDE proteins for cellular components related to EVs. Specifically, > 280 (60%) of the identified proteins mapped to GO terms extracellular membrane-bounded organelle (GO:0065010), extracellular exosome (GO:0070062), or extracellular vesicle (GO:1903561), with a > 5.7-fold (each FDR p < 2.7E-141) enrichment for each of these terms. Additional enrichment terms of interest relevant to the biogenesis of EVs were cell surface (GO:0009986) – 99 proteins, 4.6-fold enrichment, FDR p = 3E-33; and endoplasmic reticulum lumen (GO:0005788) – 59 proteins, 8.1-fold enrichment, FDR p = 1.6E-30. Classical EV markers like CD9, CD59 and multiple heat shock proteins were also identified. Relevant to the CNS, GO terms that surpassed the FDR threshold included neuronal cell body (GO:0043025) – 26 proteins, 2.2-fold enrichment, FDR p = 0.006 and axon (GO:0030424) - 28 proteins, 1.8-fold enrichment, FDR p = 0.0498. Additionally, in the human brain proteome database^28^, 19 (4.1%) and 68 (14.6%) proteins were identified as brain-enriched and brain-elevated respectively. Highly specific neuronal proteins like NCAM1/2, L1CAM, APP, NRCAM, NPTX1, NTM, VGF, NFASC, NRXN1/2/3, NPTXR, OPCML, SERPINI1, LYNX1 and NRN1 were detected in the enriched NDEs

**Table 1:** Top 20 gene ontology - cellular component enrichment terms for detected EV proteins.

| GO cellular component | Reference list (20595) | EV proteins (480) | expected | Expression | fold Enrichment | raw P-value | FDR |
| --- | --- | --- | --- | --- | --- | --- | --- |
| extracellular region (GO:0005576) | 4373 | 424 | 101.92 | + | 4.16 | 1E-211 | 2.1E-208 |
| extracellular space (GO:0005615) | 3392 | 393 | 79.06 | + | 4.97 | 8.9E-211 | 8.8E-208 |
| **extracellular membrane-bounded organelle (GO:0065010)** | 2120 | 284 | 49.41 | + | 5.75 | 7.6E-145 | 3.8E-142 |
| **extracellular organelle (GO:0043230)** | 2120 | 284 | 49.41 | + | 5.75 | 7.6E-145 | 5E-142 |
| **extracellular exosome (GO:0070062)** | 2098 | 282 | 48.9 | + | 5.77 | 7.5E-144 | 2.5E-141 |
| **extracellular vesicle (GO:1903561)** | 2118 | 283 | 49.36 | + | 5.73 | 6.7E-144 | 2.7E-141 |
| **vesicle (GO:0031982)** | 3932 | 318 | 91.64 | + | 3.47 | 5.2E-109 | 1.5E-106 |
| blood microparticle (GO:0072562) | 143 | 89 | 3.33 | + | 26.7 | 5.88E-86 | 1.46E-83 |
| collagen-containing extracellular matrix (GO:0062023) | 426 | 106 | 9.93 | + | 10.68 | 1.72E-69 | 3.8E-67 |
| extracellular matrix (GO:0031012) | 570 | 115 | 13.28 | + | 8.66 | 4.34E-67 | 8.61E-65 |
| **external encapsulating structure (GO:0030312)** | 571 | 115 | 13.31 | + | 8.64 | 5.12E-67 | 9.24E-65 |
| **vesicle lumen (GO:0031983)** | 326 | 80 | 7.6 | + | 10.53 | 1.33E-51 | 2.21E-49 |
| secretory granule lumen (GO:0034774) | 321 | 79 | 7.48 | + | 10.56 | 5.01E-51 | 7.66E-49 |
| cytoplasmic vesicle lumen (GO:0060205) | 324 | 79 | 7.55 | + | 10.46 | 9.12E-51 | 1.29E-48 |
| secretory granule (GO:0030141) | 869 | 115 | 20.25 | + | 5.68 | 6.09E-50 | 8.07E-48 |
| cell periphery (GO:0071944) | 6364 | 305 | 148.32 | + | 2.06 | 1.15E-47 | 1.43E-45 |
| **secretory vesicle (GO:0099503)** | 1037 | 116 | 24.17 | + | 4.8 | 1.1E-43 | 1.29E-41 |
| immunoglobulin complex (GO:0019814) | 189 | 56 | 4.4 | + | 12.71 | 8.49E-40 | 9.37E-38 |
| cell surface (GO:0009986) | 931 | 99 | 21.7 | + | 4.56 | 2.86E-35 | 2.99E-33 |
| **endoplasmic reticulum lumen (GO:0005788)** | 313 | 59 | 7.29 | + | 8.09 | 1.58E-32 | 1.57E-30 |

**\*In bold are terms relevant to EVs and EV biogenesis**

Among the 4 NDE preparation methods – each method that involved an additional purification step viz. M2, M3 and M4 outperformed M1 (routine NDE extraction by Exosort™) with improved protein identity parameters including protein probability, spectral counts, and unique peptide counts for brain enriched and brain elevated proteins. There were negligible differences among the three methods across the same parameters for identifying brain-enriched and brain-elevated proteins (Figure 1).

**Figure 1.**
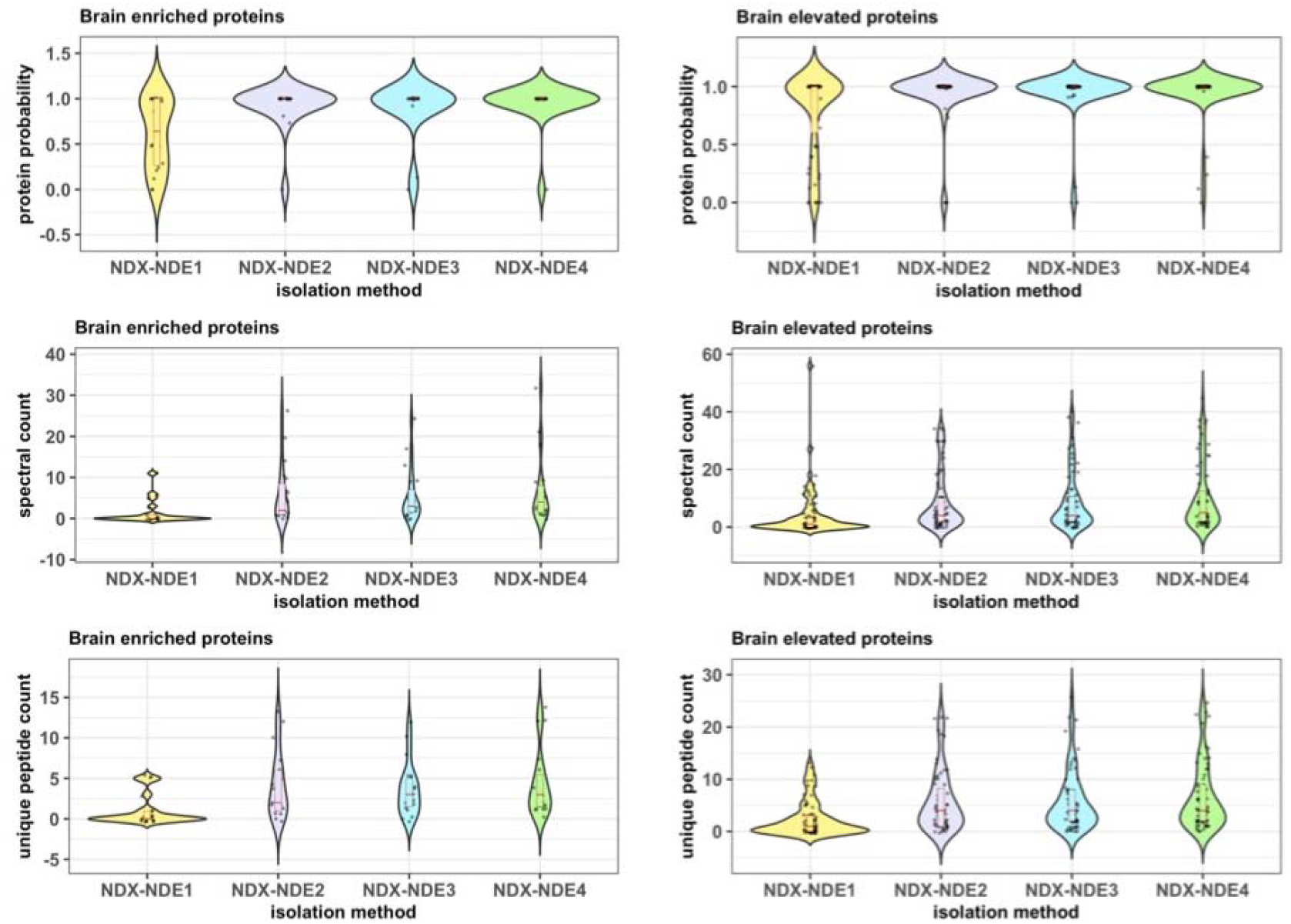
Violin plots showing the distribution density of MS parameters protein probability, spectral counts, and unique peptide counts for brain enriched and brain elevated proteins in Neuron Derived Extracellular vesicle extracts with the four different preparation methods - 1. M1 (NDX-NDE1) - Standard Exosort™ NDE extraction; 2. M2 (NDX-NDE2) - Standard Exosort™ NDE extraction + plasma protein depletion; 3. M3 (NDX-NDE3) - Erythrocyte derived EVs (CD235A, glycophorin A) depletion followed by Exosort™ NDE extraction; 4. M4 (NDX-NDE4) - Erythrocyte derived EVs depletion (CD235A, glycophorin A) followed by Exosort™ NDE extraction + plasma protein depletion. The latter three methods M2, M3 and M4 demonstrate superior protein identity parameters compared to M1. Addition of Erythrocyte EV depletion in M3 and M4 methods do not confer additional benefits when compared to plasma protein depletion in M2.

In summary, LFQ MS-proteomic analysis of Exosort based NDE preparations with additional purification steps in pooled plasma sample replicates enabled the identification of several EV relevant proteins and proteins of interest relevant to CNS. Based on these results, for the LFQ analysis of CUD and control samples, the M2 method (Exosort NDE extraction followed by depletion of 2 most abundant plasma proteins) was selected for preparing NDE lysates.

### B. Differential NDE protein abundance between CUD and controls

#### B.1 Sample profile

The sample consisted of 10 (4 females) individuals with CUD and an equal number of age and sex matched controls without CUD. The mean (SD) age of initiation of cannabis use was 15.6 (2.4) years and the mean (SD) duration of cannabis use was 6.9 (2.7) years. The mean (SD) age of the CUD and the control samples were 22.5 (1.27) and 22.9 (1.29) years respectively. The mean (SD) cannabis use in the CUD group was 47 (27) ∼number of joints in the past 30 days and 822 (668) ∼number of joints lifetime. Seven of the 10 controls had minimal lifetime exposure to cannabis but none of these participants reported having used cannabis > 2 times/week in their lifetime. None of the participants in the CUD or the control group had current or lifetime diagnosis of a major mental illness or were exposed to chronic psychotropic medications.

#### B.2 NDE proteome

A total of 231 (+/- 10) unique proteins were identified across the 20 samples from CUD and controls in the LFQ analysis. Similar to the optimization step, the enrichment profile of the identified GO cellular component terms were identical to the pilot analysis as > 153 (60.2%) of proteins mapped to GO terms extracellular membrane-bounded organelle (GO:0065010), extracellular exosome (GO:0070062), extracellular vesicle (GO:1903561) with a > 5.6-fold (FDR p < 1.2E-75) enrichment for each of these terms (Table 2). Twenty-six proteins were noted to have decreased abundance while 2 proteins had increased abundance in the CUD group compared to the control group (Table 2). The difference in the abundance of a single protein properdin encoded by the *CFP* gene between CUD and controls surpassed the significance threshold after FDR correction (fold-change = -34.9, FDR p = 0.02). Notably, SH3 and multiple ankyrin repeat domains protein 1, an adapter protein at the post-synaptic density encoded by the *SHANK1* gene was found to be depleted in the CUD compared to control NDE preparations (fold-change = -3.5, p = 0.002). Top overrepresented pathways included LXR/RXR activation (FDR p =2.00E-13), FXR/RXR activation (FDR p=1.26E-11), Coagulation system (FDR p=6.21E-07), Complement system (FDR p=2.57E-03), and a group of macrophage-related pathways among others (Supplementary Figure 2).

**Table 2:** Differentially abundant proteins in NDEs of CUD and matched controls.

| **Gene Symbol** | **Fold change** | **p-value** | **FDR p-value** | **Entrez Gene Name** |
| --- | --- | --- | --- | --- |
| CFP | -34.896 | 9.310E-05 | 2.080E-02 | complement factor properdin |
| C4A | -4.112 | 1.500E-03 | 1.460E-01 | complement C4A (Rodgers blood group) |
| IGKV3D-7 | -1.595 | 2.580E-03 | 1.460E-01 | immunoglobulin kappa variable 3D-7 |
| SHANK1 | -3.505 | 2.630E-03 | 1.460E-01 | SH3 and multiple ankyrin repeat domains 1 |
| TTR | -6.155 | 6.330E-03 | 1.880E-01 | transthyretin |
| APOA4 | -5.342 | 6.520E-03 | 1.880E-01 | apolipoprotein A4 |
| APOD | -3.711 | 6.730E-03 | 1.880E-01 | apolipoprotein D |
| FGA | -5.137 | 1.210E-02 | 3.000E-01 | fibrinogen alpha chain |
| C3 | -3.521 | 1.640E-02 | 3.470E-01 | complement C3 |
| APOA2 | -9.823 | 1.970E-02 | 3.470E-01 | apolipoprotein A2 |
| CTSD | -2.923 | 2.170E-02 | 3.470E-01 | cathepsin D |
| A2M | -2.033 | 2.210E-02 | 3.470E-01 | alpha-2-macroglobulin |
| TFPI | 2.385 | 2.430E-02 | 3.470E-01 | tissue factor pathway inhibitor |
| KLK7 | -3.648 | 2.740E-02 | 3.470E-01 | kallikrein related peptidase 7 |
| LPA | -4.855 | 2.880E-02 | 3.470E-01 | lipoprotein(a) |
| APOL1 | -3.709 | 3.230E-02 | 3.470E-01 | apolipoprotein L1 |
| RPLP2 | -33.016 | 3.360E-02 | 3.470E-01 | ribosomal protein lateral stalk subunit P2 |
| LCN1 | -3.855 | 3.460E-02 | 3.470E-01 | lipocalin 1 |
| CSTA | -3.317 | 3.490E-02 | 3.470E-01 | cystatin A |
| ENO1 | -2.239 | 3.620E-02 | 3.470E-01 | enolase 1 |
| PRDX2 | -34.5 | 3.730E-02 | 3.470E-01 | peroxiredoxin 2 |
| GGCT | -2.817 | 3.760E-02 | 3.470E-01 | gamma-glutamylcyclotransferase |
| HBB | -2.178 | 3.870E-02 | 3.470E-01 | hemoglobin subunit beta |
| F11 | 3.092 | 3.890E-02 | 3.470E-01 | coagulation factor XI |
| CTSB | -3.271 | 4.130E-02 | 3.540E-01 | cathepsin B |
| DCD | -2.022 | 4.840E-02 | 4.000E-01 | dermcidin |

#### B.3 Validation of proteomic signatures with ELISA

Three of the top differentially abundant proteins were validated using ELISA. CFP was measured in 8-fold dilution using ELISA kit (Thermo Scientific, Cat. EH383RB). C4A was measured in 2-fold dilution using Luminex kit (Sigma-Aldrich, Cat. HCMP2MAG-19K-07). SHANK1 was measured without any dilution using ELISA kit (MyBioSource Cat. MBS9339587) (Figure 3). A positive correlation was noted between the MS and the ELISA concentrations for each protein - CFP (spearman rho = 0.52, p = .018), C4A (spearman rho = 0.32, p = .17) and SHANK1 (spearman rho = 0.36, p = .12). Within the CUD sub-sample, a statistically significant correlation was noted between MS and ELISA concentrations for SHANK1 (spearman rho = 0.7, p = 0.02).

**Figure 2.**
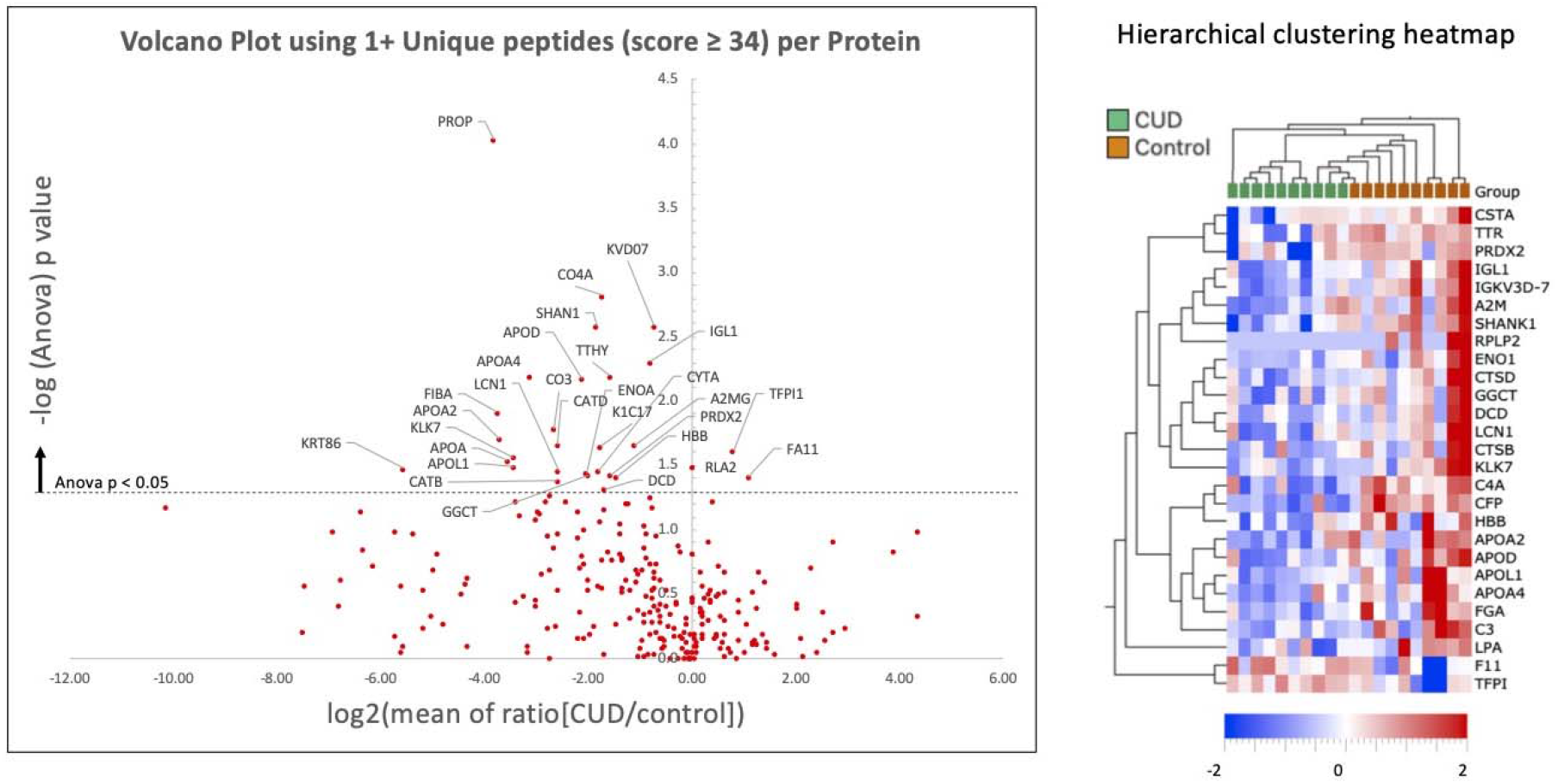
A. Volcano plot showing the differentially abundant proteins between CUD and control samples with Y axis representing the negative log of ANOVA p values and X axis representing the log 2 transformed ratios of abundance between CUD and controls. The horizontal dotted line represents a nominal significance threshold of p < 0.05. B. A hierarchical clustering heatmap showing differentially abundant proteins that surpass nominal significance threshold.

**Figure 3.**
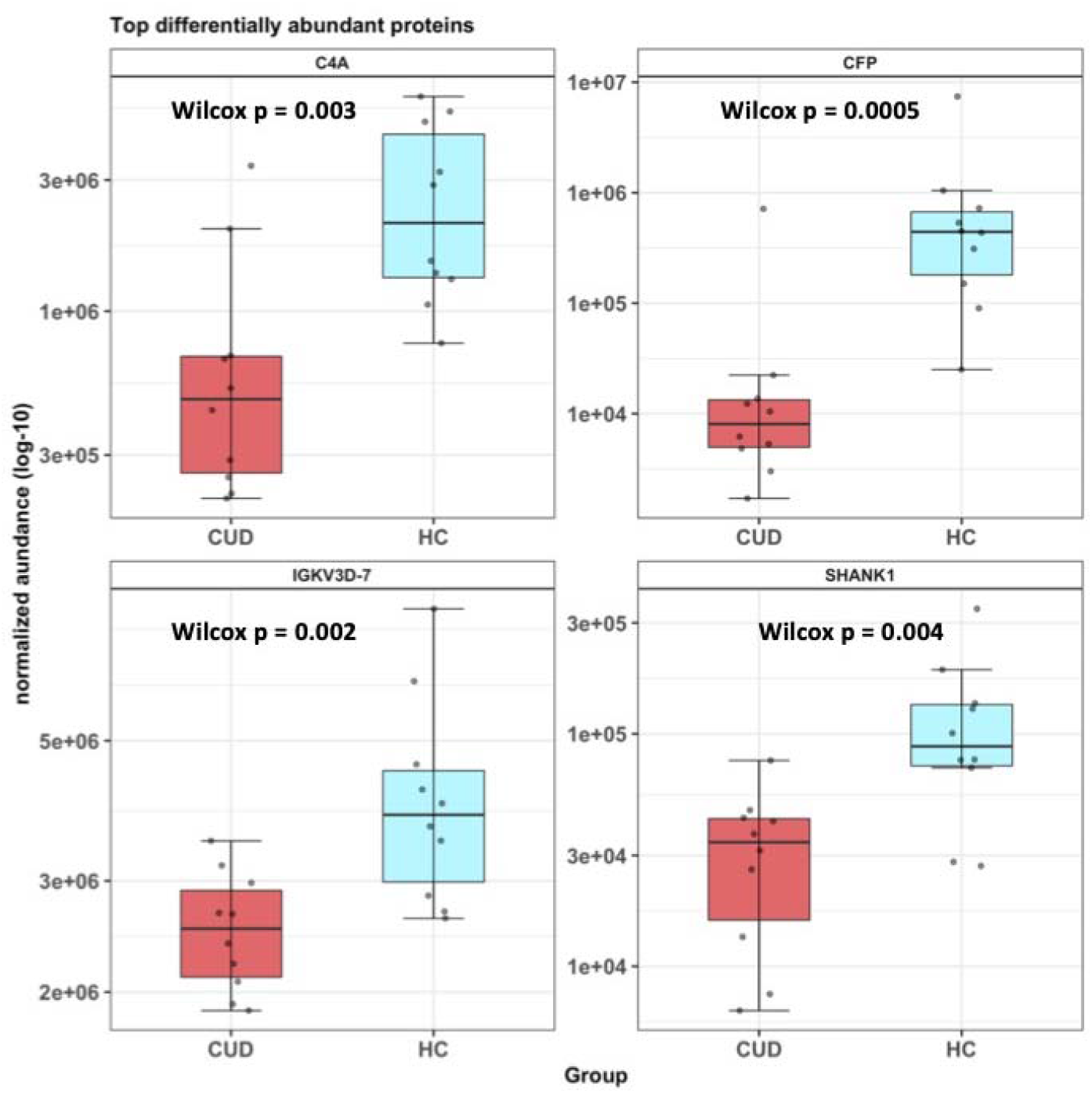
Box and Whisker plots showing the top 4 differentially abundant proteins between CUD and control NDE preparations. These proteins included CFP (Properedin), C4A (complement factor 4A), IGKV3D-7 (Immunoglobulin Kappa Variable 3D-7) and SHANK1 (SH3 and multiple ankyrin repeat domains protein 1). Y axis represents log transformed normalized abundance values.

## Discussion

To our knowledge, this study represents one of the first attempts to employ a discovery proteomics approach to characterize the proteome of plasma NDEs to identify peripheral signatures of neuropathology in young onset CUD. There is considerable interest in the application of EV assays to discover novel disease relevant biomarkers in neuropsychiatric syndromes ^29^. Here we successfully demonstrate feasibility and utility of LFQ proteomic profiling of plasma NDEs to identify protein biomarkers that suggest potential underlying neuropathology secondary to chronic recurrent cannabis exposure.

### Optimal methods to identify protein biomarkers in NDE

Several studies to date have employed discovery MS proteomic approaches to identify protein biomarkers in plasma derived EVs in neurodegenerative disorders and in traumatic brain injury. However, we were able to find a single study in the literature by Anastasi et al, that has employed MS proteomics on plasma NDEs in a single subject with Parkinson’s disease ^30^. Using a LC-MS/MS approach, the authors were able to identify ∼349 protein groups of which 20 proteins were annotated as brain elevated in Human Protein Atlas. In the present study, we identified a higher number of total proteins (465) and brain expression elevated proteins (68) from pooled plasma replicates during protocol development.

As noted in a recent review, variability in EV isolation due to non-exosomal vesicle contamination, co-isolation of protein aggregates and lipoproteins, and vesicle membrane damage also pose challenges to application of sensitive MS proteomic approaches to study plasma NDEs ^29^. In this study, we note that ∼40% of identified proteins are non-EV proteins. It is yet unclear if these represent technical contamination or proteins that interact with EVs, it is well know that both EVs and several abundant plasma proteins are highly sticky (PMC6208672, PMC4834552). However, with the depletion of two most abundant plasma proteins after NDE enrichment with Exosort, we were able to identify several brain-expressed proteins in plasma NDE isolates. The addition of blood microparticle depletion step prior to NDE enrichment resulted in comparable protein identity parameters and hence was not adopted for further analysis.

### Relevance of NDE protein biomarkers to CUD

We noted 28 differentially abundant proteins with 26 proteins having decreased abundance and 2 having increased abundance in CUD compared to controls NDE proteome. SHANK1, an adapter protein involved in the structural and functional integrity of glutamatergic post-synapses, was among the four most differentially abundant proteins. SHANK1 is abundantly and almost exclusively expressed in the brain and is enriched in glutamatergic synapses in the cortex, thalamus, amygdala, hippocampus, dentate gyrus and cerebellar Purkinje neurons ^31^. The SHANK family of proteins interact directly with multiple post synaptic density proteins and indirectly with NMDA, AMPA and kainite type glutamatergic receptors (Supplementary Figure 3, SHANK1 interactome) thus playing an important role in synapse formation, maturation, and synaptic plasticity ^32^. SHANK1 knock out mice demonstrated rapid disintegration of post synaptic densities ^32^.

Chronic exposure to major phyto-cannabinoids such as delta-9-tetrahydrocannabinol (THC), Cannabidiol (CBD), and Cannabigerol (CBG) has been found to affect expression of synaptic proteins in both preclinical and in vitro cellular models. Chronic THC exposure in human induced pluripotent stem cell (hiPSC) derived neurons was noted to alter the expression of multiple post synaptic density proteins ^33^. Interestingly this study noted > 4-fold reduction in the expression of SHANK1 in hiPSC neurons following chronic but not acute exposure to THC. Similarly, varying concentrations of both CBD and CBG have also been reported to downregulate the expression of SHANK1 over 24 hours in NSC-34 motor neuron like cells ^34^. Chronic THC exposure in adolescent female rats has been demonstrated to result in downregulation of expression of SHANK1 interacting PSD-95 and synaptophysin proteins in the prefrontal cortex and hippocampus ^35^. Multiple lines of evidence support alterations in synaptic plasticity and neuronal wiring following recurrent exposure to cannabis during neurodevelopment possibly mediated via dysregulated expression of synaptic proteins ^36^. Using Positron Emission Tomography, we have previously demonstrated reduction in synaptic vesicle density in CUD specifically with large effects in the hippocampus ^37^. Taken together, the finding of reduced SHANK1 abundance in plasma NDE proteome in CUD may suggest a novel peripheral biomarker for altered brain signalling in glutamatergic synapses secondary to cannabis exposure.

Among the differentially abundant proteins we noted several proteins of the complement and coagulation cascade. Both complement proteins and coagulation cascade proteins are expressed in the brain and altered expression of these proteins is reported in complex CNS syndromes ^38, 39^. However, unlike SHANK1 protein with a relatively high abundance in the brain, these proteins are also abundant in plasma. Hence the precise origin of these proteins and the significance of the differential abundance to CUD cannot be fully determined from the present study. Interestingly, the top overrepresented canonical pathways among differentially abundant proteins identified in the present study that included LXR/RXR activation, FXR/RXR activation, acute phase response, coagulation and complement pathways, replicated a previous analysis differentially abundant proteins in serum proteome between CUD and controls ^40^.

### Strengths, limitations, future directions

The results are to be interpreted in the background of some limitations. With respect to the purity of the EV preparations, using a LFQ proteomic approach we were able to detect ∼ 60% of the proteins previously annotated as EV related proteins. However, the remaining fraction may be contributed by residual plasma contamination. Further efforts are necessary to optimize techniques of EV isolation, enrichment and purification to enhance the signal to noise ratio to detect proteins of interest using LFQ analysis. We detected a relatively larger number of proteins in the pooled plasma samples compared to the CUD-control samples. This could be resulting from a heterogenous population of EVs in the pooled plasma.

In conclusion, we demonstrate that a label-free quantitative MS proteomics approach to analyze NDEs enriched from plasma may yield significant insights into the synaptic pathology underlying CUD. Additional optimization of these methods could result in a novel assay to study altered signalling in the brain using liquid biopsy across diverse neuropsychiatric syndromes.

## Data Availability

All data produced in the present study are available upon reasonable request to the authors

## Funding

SG is supported by NARSAD young investigator award #27340 from the Brain and Behavior Research Foundation to study longitudinal effects of cannabis exposure in adolescents and young adults. The present work was additionally supported by a pilot award to study altered brain proteomic signalling in addiction by the Yale/NIDA Neuroproteomic Center (DA018343). ACN and T.T.L were also supported in part by the Yale/NIDA Neuroproteomics Center (DA018343). The Q-Exactive HFX mass spectrometer and M-class UPLC at the Keck MS & Proteomics Resource was supported in part by NIH SIG grants OD023651-01A1 and the Yale School of Medicine. The funders had no role in study design, data collection and analysis, decision to publish, or preparation of the manuscript.

We also like to thank Florine Collin and Weiwei Wang from the Keck MS & Proteomics Resource for their support on sample preparation and data collection, respectively.

