## supplementary table1 for "Peripheral signature of altered synaptic integrity in young onset Cannabis Use Disorder: A proteomic study of circulating extracellular vesicles"

Supplementary table 1 – Overrepresentation analysis of all proteins identified across CUD and matched control samples

| **GO cellular component complete** | **Homo sapiens - REFLIST (20595)** | **Input (267)** | **expected** | **over/under** | **fold Enrichment** | **raw P-value** | **FDR** |
| --- | --- | --- | --- | --- | --- | --- | --- |
| extracellular space (GO:0005615) | 3386 | 208 | 43.9 | + | 4.74 | 2.35E-106 | 4.67E-103 |
| extracellular region (GO:0005576) | 4370 | 219 | 56.65 | + | 3.87 | 4.63E-98 | 4.60E-95 |
| **extracellular membrane-bounded organelle (GO:0065010)** | **2120** | **155** | **27.48** | **+** | **5.64** | **1.19E-79** | **5.93E-77** |
| **extracellular organelle (GO:0043230)** | **2120** | **155** | **27.48** | **+** | **5.64** | **1.19E-79** | **7.90E-77** |
| **extracellular vesicle (GO:1903561)** | **2118** | **154** | **27.46** | **+** | **5.61** | **1.18E-78** | **4.68E-76** |
| **extracellular exosome (GO:0070062)** | **2098** | **153** | **27.2** | **+** | **5.63** | **3.63E-78** | **1.20E-75** |
| vesicle (GO:0031982) | 3930 | 176 | 50.95 | + | 3.45 | 1.74E-61 | 4.93E-59 |
| blood microparticle (GO:0072562) | 143 | 49 | 1.85 | + | 26.43 | 5.92E-50 | 1.47E-47 |
| vesicle lumen (GO:0031983) | 327 | 56 | 4.24 | + | 13.21 | 7.43E-43 | 1.64E-40 |
| secretory granule lumen (GO:0034774) | 321 | 55 | 4.16 | + | 13.22 | 4.37E-42 | 8.69E-40 |
| cytoplasmic vesicle lumen (GO:0060205) | 325 | 55 | 4.21 | + | 13.05 | 7.86E-42 | 1.42E-39 |
| secretory granule (GO:0030141) | 867 | 77 | 11.24 | + | 6.85 | 1.16E-40 | 1.91E-38 |
| secretory vesicle (GO:0099503) | 1035 | 78 | 13.42 | + | 5.81 | 1.58E-36 | 2.42E-34 |
| collagen-containing extracellular matrix (GO:0062023) | 426 | 51 | 5.52 | + | 9.23 | 3.73E-32 | 5.30E-30 |
| extracellular matrix (GO:0031012) | 571 | 54 | 7.4 | + | 7.29 | 2.34E-29 | 3.10E-27 |
| external encapsulating structure (GO:0030312) | 572 | 54 | 7.42 | + | 7.28 | 2.53E-29 | 3.15E-27 |
| intermediate filament (GO:0005882) | 228 | 37 | 2.96 | + | 12.52 | 1.39E-27 | 1.63E-25 |
| cytoplasmic vesicle (GO:0031410) | 2450 | 100 | 31.76 | + | 3.15 | 2.08E-26 | 2.30E-24 |
| intermediate filament cytoskeleton (GO:0045111) | 269 | 38 | 3.49 | + | 10.9 | 2.37E-26 | 2.48E-24 |
| intracellular vesicle (GO:0097708) | 2456 | 100 | 31.84 | + | 3.14 | 2.50E-26 | 2.49E-24 |

* In bold are GO terms relevant to EVs and EV biogenesis

Supplementary figure 1 NDE isolation workflow, electron microscopy and nanoparticle tracking analysis

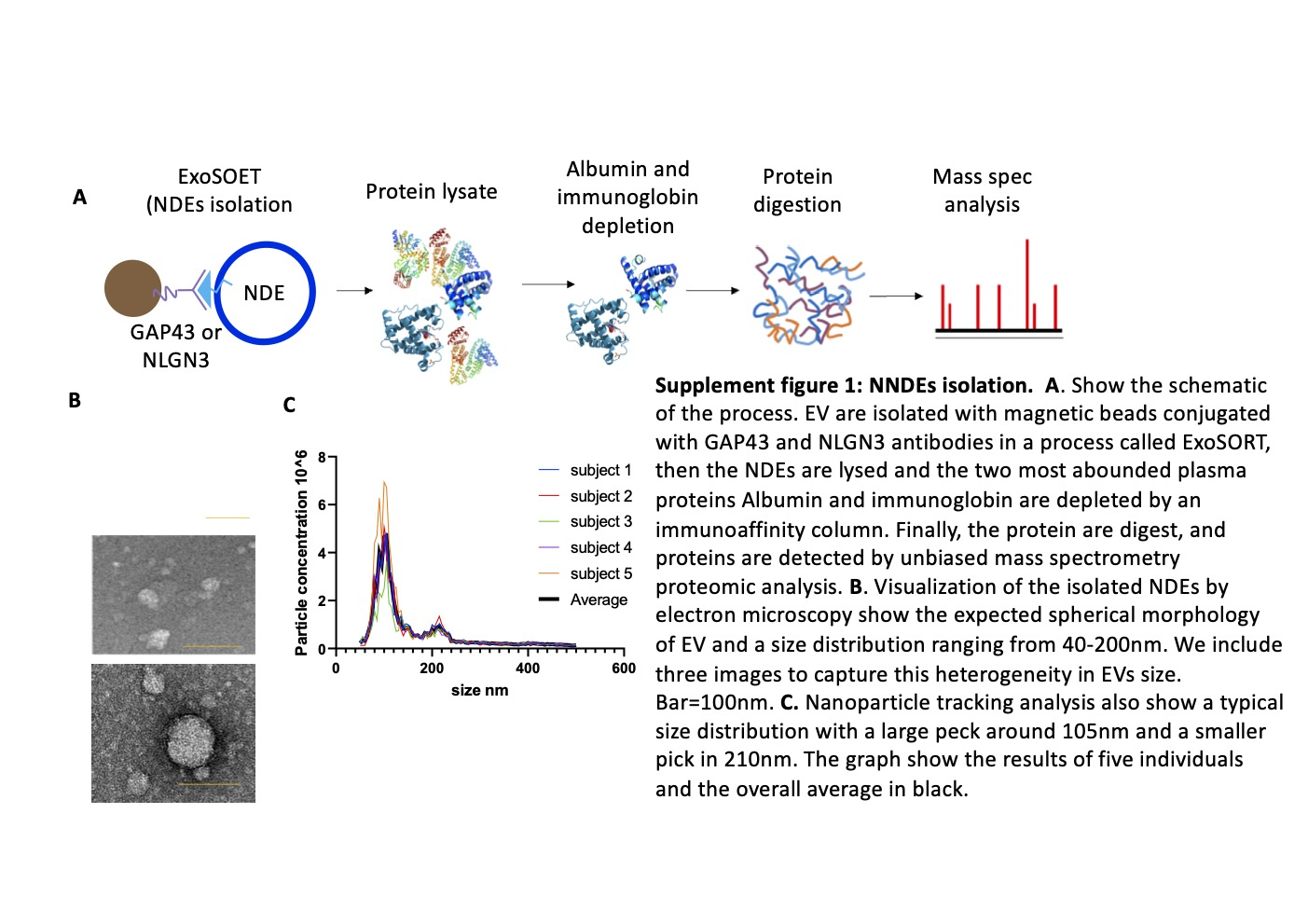

Supplementary figure 2 – Overrepresentation analysis of canonical pathways for differentially abundant proteins between CUD and controls

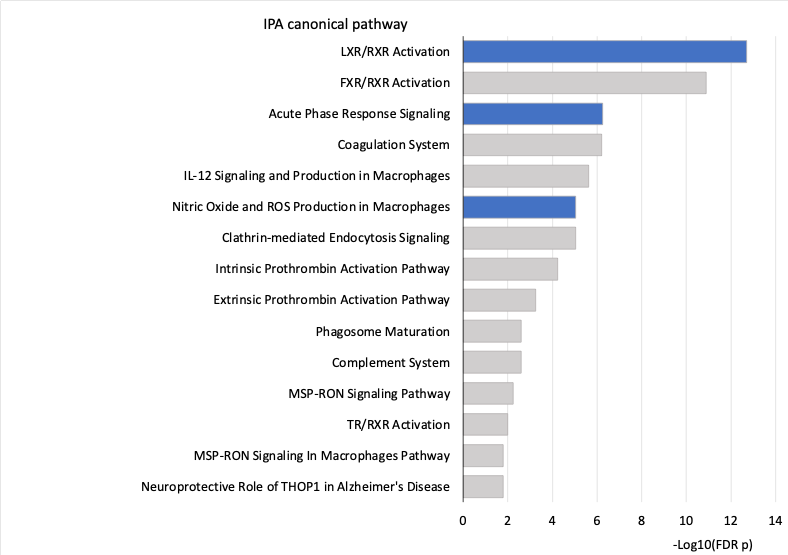

Supplementary figure 3 – SHANK1 protein interactome demonstrating interaction with multiple metabotropic and ionotropic glutamate receptor proteins

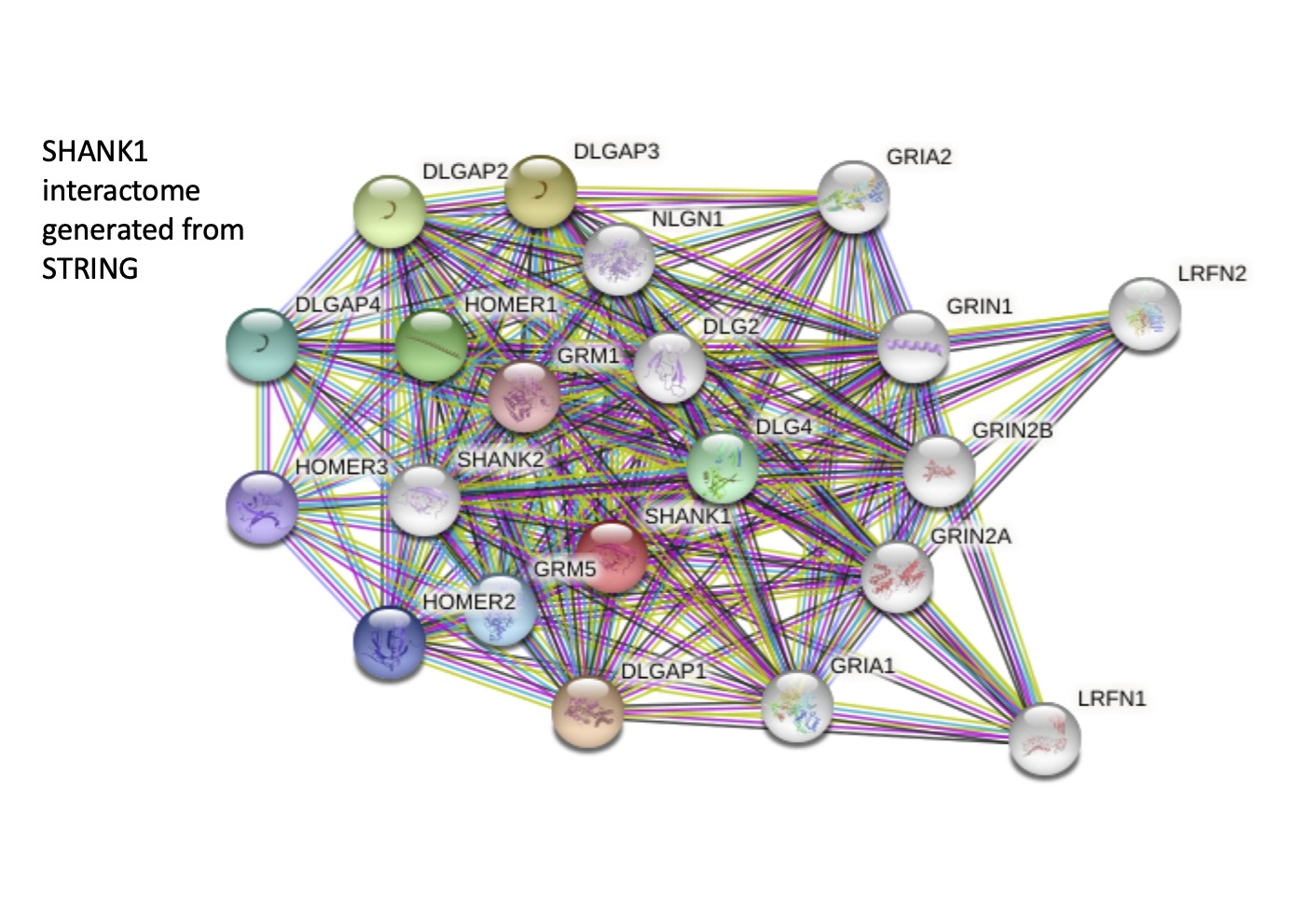
